# The impact of remote consultations on the quality of primary care: A systematic review

**DOI:** 10.1101/2023.05.05.23289593

**Authors:** Kate Campbell, Geva Greenfield, Edmond Li, Niki O’Brien, Benedict Hayhoe, Thomas Beaney, Azeem Majeed, Ana Luisa Neves

**Author notes:** **Corresponding author:** Dr Ana Luisa Neves^1^, Department of Primary Care and Public Health, Imperial College London.

## Abstract

**Background:** The adoption of remote consultations, catalysed by the COVID-19 pandemic, has transformed the delivery of primary care services. We evaluated the impact of remote consultations on the quality of primary care.

**Methods:** Six databases were searched. Studies evaluating the impact of remote consultations, for any disease, were included. Title and abstract screening, and full-text screening were performed by two pairs of investigators. Risk of bias was assessed using the Mixed Methods Appraisal Tool. A narrative synthesis of the results was performed.

**Findings:** Thirty studies (5,469,333 participants) were included in the review. Remote consultations generally had a positive or equivalent impact compared to face-to-face (F2F) consultations, particularly in reducing patient costs and improving time efficiency. The effectiveness of remote consultations was non-inferior to F2F care in six out of seven studies evaluating this aspect. Two studies found that remote consultations reduced wait times for appointments. Younger, female patients were more likely to use remote consultations and those of lower socioeconomic status were less likely to use video consultations than telephone appointments. The impacts on safety and patient-centeredness were largely inconclusive.

**Interpretation:** Remote consultations may be equally as effective as F2F care and have a potentially positive impact on the efficiency and timeliness of care. Those of lower socioeconomic status were more likely to use consultations delivered via telephone than videoconference. Developing a strong evidence-base capitalising on real-world data as well as clinical trials is crucial for the future development of remote consultations and tailoring them to patient needs and preferences.

**Funding:** National Institute for Health and Care Research Applied Research Collaboration Northwest London.

**What is already known on this topic:** Existing literature reviews exploring remote consultations have primarily been confined to assessing their impact on effectiveness, efficiency, or specific clinical conditions whilst utilising a broad definition regarding what constitutes remote services. Evidence was largely heterogeneous, often focussing on interventions delivered in secondary care facilities or by specialists only. There is a paucity of systematic reviews pertaining to primary care.

**What this study adds:** This systematic review investigates the impact of remote consultations on the quality of primary care. Our results show that remote consultations may be equally as effective as F2F care and have a potentially positive impact on efficiency, timeliness of care, and reduced rates of follow-up in secondary or tertiary care. Patients from lower socioeconomic backgrounds were more likely to use consultations delivered via telephone than video conference.

**How this study might affect research, practice or policy:** Our systematic review has demonstrated that remote consultations have the potential to be just as effective as F2F consultations by reducing waiting times, patient costs, and rates of follow-up in hospitals. However, there currently remains a lack of robust studies available exploring the effect of remote consultations on patient safety, equity, and patient-centredness, highlighting areas where future research efforts need to be devoted. Data collection methods more bespoke to the primary care context, better accounting for patient characteristics and needs, and inclusive of its intended end-users, are necessary to generate a stronger evidence base to inform future remote care policies.

## Introduction

The onset of the COVID-19 pandemic in 2020 resulted in the rapid expansion of remote consultations in primary care.^1^ The shift from a primarily face-to-face (F2F) model of healthcare provision led to approximately 70%^2^ and 65%^3^ of primary care contacts being delivered remotely in the United Kingdom (UK) and the United States (US), respectively.

Remote consultations can be defined as real-time communication between patients and clinicians, through telephone or videoconferencing.^4^ It is argued that not only have remote consultations helped to minimise COVID-19 transmission, but they may also improve efficiency and access to care.^5^ This may be particularly important in rural areas with geographical disparities in service provision^6^ and resource-constrained settings with workforce shortages.^4^

However, concerns have been raised over the speed at which remote consultations were implemented, with both patients and clinicians reporting a lack of confidence in the underlying technology and potentially poorer clinical decision-making as key issues.^7^ Remote care limits clinicians’ capacity to conduct physical examinations^8^ and increase reliance on patients’ abilities to articulate their symptoms,^5^ posing potential safety risks. Moreover, those with limited access to technology or with lower digital literacy may be at risk of ‘digital exclusion’.^9^

Previous reviews have investigated the impact of remote consultations on the effectiveness^3,10-13^ and efficiency^14,15^ of care, often focussing on specific clinical conditions or taking broad definitions of remote services. However, there is a notable lack of systematic reviews assessing the impacts on safety, patient-centredness, timeliness and equity, with much of the existing literature limited to scoping or rapid reviews of the evidence.^16-19^ Furthermore, while some systematic reviews investigate the impact of remote care on aspects of quality exclusively in primary care settings,^11,14,15,20^ others include heterogeneous evidence, including interventions delivered in secondary care facilities or by specialists.^3,13^ There is therefore a need to comprehensively evaluate the impact on all aspects of care quality, in this specific clinical setting.

The aim of this review is to systematically evaluate the impact of remote consultations on the quality of primary care. We chose to use the Institute of Medicine (IOM)’s theoretical framework to map the impact across six domains of quality, including efficiency, effectiveness, safety, patient-centredness, timeliness and equity.^21^

## Methods

This systematic review followed the Preferred Reporting Items for Systematic Review and Meta-Analysis (PRISMA) checklist.^22^ The study protocol was registered with the International Prospective Register of Systematic Reviews (CRD42022362380).

### Search Strategy

Six databases (MEDLINE, Embase, HMIC, PsychInfo, CINAHL and Cochrane) were searched on 20^th^ June 2022. The search included studies published between January 2017 to June 2022, as the last 5 years have seen most of the shift toward remote care. A combination of free text and Medical Subject Headings (MeSH) terms was used (Appendix 1). The concepts of remote consultations and primary care were kept intentionally broad to address variations in language; search terms for the domains of quality were adapted from a previous review.^23^ Reference lists of relevant systematic reviews and grey literature sources were also screened.

### Study Selection Criteria

Studies were included if they focused on adult patients accessing primary care services; involved telephone or videoconference consultations delivered by healthcare professionals; compared outcomes with F2F consultations; and reported outcomes that fit under any of the IOM’s quality of care domains. A detailed description of inclusion and exclusion criteria is provided in Appendix 2. Studies focussing on specific health conditions were not excluded, in order to characterise the general use of remote consultations in primary care.

### Screening and Data Extraction

Following the removal of duplicates, citations were uploaded into an online screening tool (Covidence^24^). Title and abstract screening, followed by full-text screening, were performed by two independent reviewers. Cohen’s kappa was used to measure intercoder agreement in each screening phase (0.22 and 0.65, respectively). Disagreements were resolved by discussion with a third investigator. Data extraction was conducted using a standardised data extraction form (Appendix 3). Effect sizes such as mean differences, odds ratios (OR) and risk ratios (RR) were extracted. Where available, rates of intervention adherence, follow-up and survey response were also extracted.

### Risk of Bias Assessment

Risk of bias was assessed using the Mixed Methods Appraisal Tool (MMAT v.18)^25^ (Appendix 4). A study was considered high risk if it scored ‘Yes’ in two or fewer dimensions, moderate risk if it scored ‘Yes’ in three dimensions, and low risk if it scored ‘Yes’ in four or all dimensions.

### Data Synthesis

A narrative synthesis of the findings was conducted for each domain of quality.

### Role of the funding source

The study funders did not play a role in the study design; collection, analysis, or interpretation of data; manuscript writing; or in the decision to submit for publication. Researchers were independent of the funders. All authors had full access to all data included in this study and can take responsibility for its integrity and the accuracy of the data analysis.

## Results

### Search Results

Searches retrieved a total of 6,272 records (Figure 1). No relevant records were found from searching grey literature or from reference lists of relevant articles. Thirty papers met the inclusion criteria.

**Figure 1.**
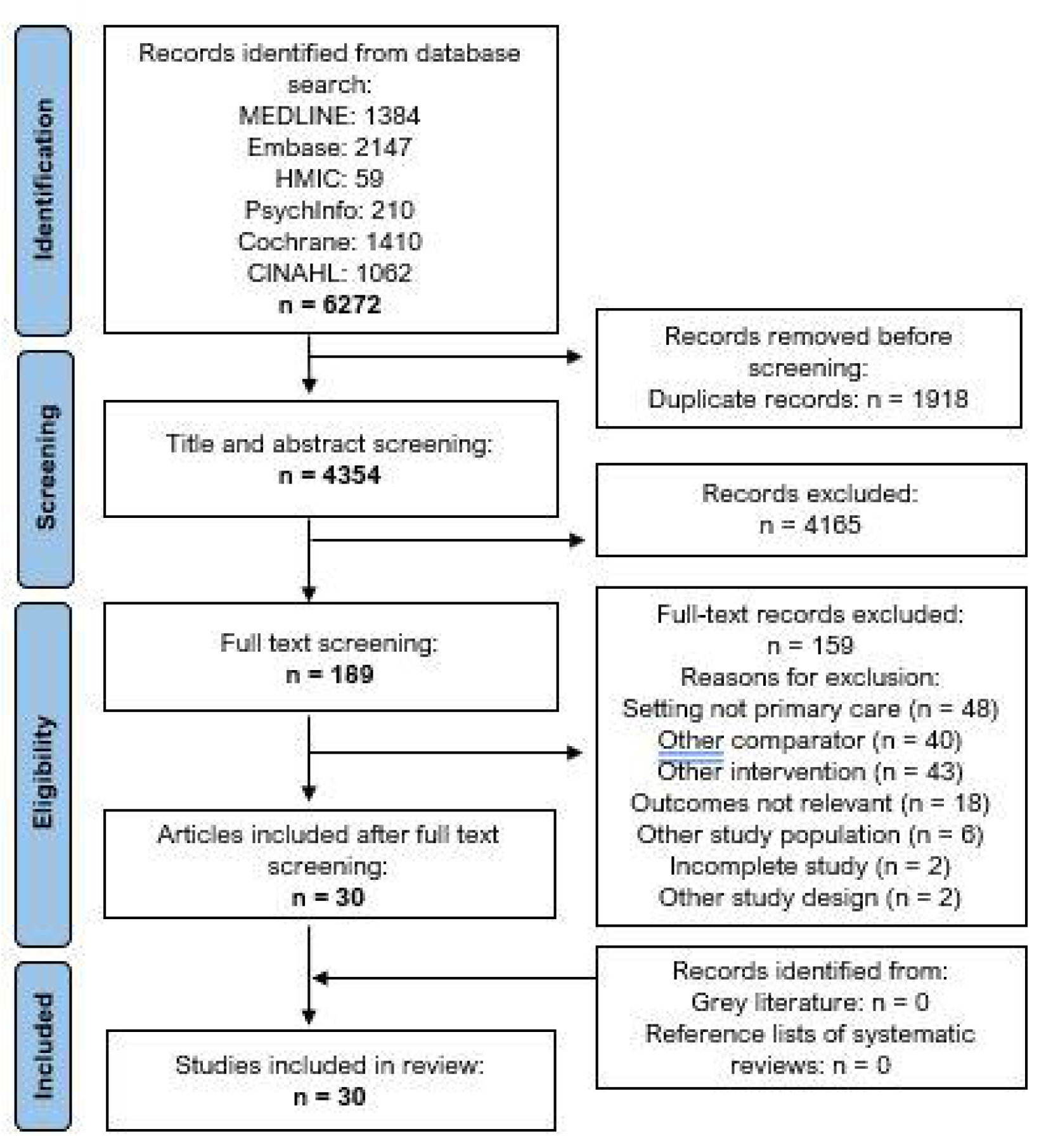
PRISMA flow chart of included studies.

### Description of Included Studies

The 30 included studies (Table 1) comprised a total of 5,469,333 subjects. Sample sizes ranged from 28^26^ to 1,490,734^27^ participants, and publication years ranged between 2017 and 2022. Study types included 14 retrospective cohorts,^27-40^ six cross-sectional,^41-46^ four quasi-experimental,^26,47-49^ three randomised controlled trials (RCT),^50-52^ two cohorts,^53,54^ and one cluster RCT.^55^ Most studies took place in the US (n=20),^27-31,33-41,43-46,50,55^ with the rest conducted in Australia,^32,42^ Canada,^47,49^ Kenya,^51^ the UK,^48^ Japan,^52^ Singapore,^26^ New Zealand,^53^ and Sweden.^54^

**Table 1.** Characteristics of the included studies.

| Author, year | Country | Time period | Study design | Participants (setting) | Sample size (N) | Study design | Consultation description | Risk of bias |
| --- | --- | --- | --- | --- | --- | --- | --- | --- |
| **Baughman, 2022a**<sup>28</sup> | USA | April 2020 - September 2021 | Retrospective cohort study | Primary care patients aged 26-70 with abnormal BMI scores (<18.5 or >25 (kg/m<sup>2</sup>)) | 287,387(RC: 1,556; blended: 63,489; F2F: 222,333) | Comparison of outcomes between initial visit modalities:<br>RC (Zoom video)<br>blended RC (patients with RC and F2F visits within the timeframe)<br>F2F | Initial BMI screening visits | Moderate |
| **Baughman, 2022b**<sup>29</sup> | USA | July 2019 - June 2021 | Retrospective cohort study | Primary care patients aged 18-50 with a lower back pain diagnosis | 20,624 (RC: 5,334; F2F: 15,290) | Comparison of outcomes between initial visit modalities:<br>RC (Zoom video)<br>F2F | Initial primary care consultation | Moderate |
| **Befort, 2021**<sup>55</sup> | USA | February 2016 - October 2017 | Cluster RCT | Primary care patients aged 20-75 (mean age 54.7) with BMI scores of 30 - 45, residing in rural locations | 1,407 (group RC: 466; individual F2F: 473; group F2F: 468) | Practices (n=36) randomly assigned to:<br>group RC (telephone conference call)<br>group F2F<br>individual F2F | Individual F2F: 15-minute behavioural therapy;<br>Group RC and F2F: 60-minute group behavioural therapy (14 patients per group) | Moderate |
| **Bernstein, 2021**<sup>30</sup> | USA | November 2015 - March 2019 | Retrospective cohort study | Ambulatory care patients aged >60 with urgent and non-emergent conditions | 1088(RC: 115; F2F: 973) | Comparison of outcomes between index visit modalities:<br>RC (video)<br>F2F | Visit with physician including clinical assessment, prescriptions, or referrals if appropriate | Moderate |
| **Chavez, 2022**<sup>31</sup> | USA | April 2019 - March 2021 | Retrospective cohort study | Patients (mean age 54.1) accessing care at an academic family medicine practice | 57006<sup>a</sup> (RC: 7,577; F2F: 49,429) | Comparison of outcomes between index visit modalities:<br>RC (video or telephone)<br>F2F<br>Stratified by pre-pandemic and pandemic timeframes | Consultation with physician | High |
| **Dai, 2022**<sup>32</sup> | Australia | March 2020 - August 2021 | Retrospective cohort study | Geriatric primary care patients aged >65 in residential aged care facilities | 27,980 | Assessed associations between sociodemographic characteristics and visit modality:<br>RC (video or telephone)<br>F2F | GP consultation | Low |
| **Egede, 2017**<sup>50</sup> | USA | September 2006 - October 2012 | RCT | Elderly veterans aged >58 with depression | 241 (RC: 120; F2F: 121) | Comparison of outcomes between consultation modalities: | 60-minute behavioural activation for depression | Moderate |
|  |  |  |  |  |  | RC (videophone)<br>F2F | delivered weekly for 8 weeks |  |
| **Frank, 2021**<sup>33</sup> | USA | March 2019 - December 2020 | Retrospective cohort study | Mental health patients at an academic primary care practice aged 4 – 73 (mean age 28.32) | 173 | Comparison of outcomes between two waves of consultation modalities:<br>RC (video or telephone): Mar - Dec 20<br>F2F: Mar - Dec 19 | 30-minute mental health appointments | High |
| **Gordon, 2017**<sup>34</sup> | USA | January 2014 - May 2015 | Retrospective cohort study | Primary care patients aged <65 receiving care for acute, nonurgent conditions<sup>b</sup> | 18,516 (RC: 4,635; F2F: 13,881) | Comparison of outcomes between consultation modalities:<br>RC<br>F2F | Primary care consultations | Moderate |
| **Govier, 2022**<sup>35</sup> | USA | March 2019 - July 2021 | Retrospective cohort study | Primary care patients aged >18 diagnosed with COVID-19 between March - July 2020 | 11,326 (RC:1,360; F2F: 9,966) | Assessed associations between sociodemographic characteristics and visit modality:<br>RC<br>F2F | Primary care consultations | Moderate |
| **Graetz, 2022**<sup>36</sup> | USA | January 2016 - May 2018 | Retrospective cohort study | Primary care patients of all ages (22.2% >65) | 1,131,722 | Comparison of outcomes between consultation modalities: | Primary care consultations requested by patients using an | Moderate |
|  |  |  |  |  |  | RC (video)<br>RC (telephone)<br>F2F | online portal |  |
| **Haderlein, 2022<sup>37</sup>** | USA | March 2018 - October 2021 | Retrospective cohort study | Veterans (mean age 48.7) seeking mental health care in an urban VA Primary Care-Mental Health Integration clinic | 2,479 | Comparison of outcomes between consultation modalities:<br>RC (video or telephone)<br>F2F | Primary care mental health consultations | Low |
| **Harder, 2020<sup>51</sup>** | Kenya | September 2014 - December 2015 | RCT | Primary care patients (mean age 38) with alcohol use disorders in rural primary health centre | 300 (RC:104; F2F: 92; Waitlist control: 104) | Comparison of outcomes between consultation modalities:<br>RC (mobile phone call)<br>F2F<br>Waitlist control | One 30-minute motivational interviewing session | High |
| **Li, 2022<sup>27</sup>** | USA | June 2019 - September 2020 | Retrospective cohort study | Primary care patients (mean age 39.2) | 1,490,734 | Comparison of outcomes between practice level of RC use:<br>High RC use<br>Medium RC use<br>Low RC use | Primary care consultations | High |
| **Lovell, 2021**<sup>41</sup> | USA | April 2016 - March 2017 | Retrospective cohort study | Primary care patients aged <64 accessing care for low-acuity, urgent conditions<sup>c</sup> | 5,919 (RC: 1,531; F2F: 4,388) | Comparison of outcomes between consultation modalities:<br>RC (videocall)<br>F2F | Primary care consultations | High |
| **Manski-Nankervis, 2022**<sup>42</sup> | Australia | October 2020 -May 2021 | Descriptive cross-sectional study | Primary care patients (mean age 31.8) who completed a videoconference call with a health care professional | 499 | Online survey of experience with most recent RC visit (videoconference) with comparison to past experience of F2F visit | Primary care consultations | High |
| **McGrail, 2017**<sup>47</sup> | Canada | 2013 - 2014 | Quasi-experimental (interrupted time series) with cross-sectional survey component | Primary care patients aged >18 living in British Colombia | 29,267 (RC: 7,286; F2F: 21,981)<br>Survey: 399 | Comparison of outcomes between consultation modalities:<br>RC<br>F2F<br>Online survey of experience with most recent RC visit with comparison to past experience of F2F visit | Primary care consultations | Moderate |
| **Miller, 2019**<sup>48</sup> | UK | June 2014 – May 2017 | Quasi-experimental (interrupted time series) | Primary care patients of all ages at an urban general practice in a socio-economically | 27,589 (RC: 9,113; F2F: 18,476) | Comparison of outcomes between the two study phases:<br>F2F (preintervention phase) | Primary care consultations | Moderate |
|  |  |  |  | deprived area |  | RC (telephone-first phase) |  |  |
| **Mohan, 2022<sup>43</sup>** | USA | April - December 2020 | Descriptive cross-sectional study | Primary care patients (mean age 48.7) at academic medical centre | 797 | Online survey of experience with most recent RC visit with comparison to past experience of F2F visit | Primary care consultations | High |
| **Neufeld, 2022<sup>49</sup>** | Canada | September 2020 - February 2021 | Quasi-experimental study | Primary care patients aged 18-87) | 66 (RC: 32; F2F: 34) | Comparison of outcomes between consultation modalities:<br>RC (telephone)<br>F2F<br>Assessed by online survey | Family physician consultations | Moderate |
| **Nomura, 2019<sup>52</sup>** | Japan | March - June 2018 | RCT | Primary care patients (mean age 55) with nicotine dependence | 115 (RC: 58; F2F: 57) | Comparison of outcomes between consultation modalities:<br>RC (internet-based video call)<br>F2F | Smoking cessation counselling, 5 sessions over 24 weeks with access to smoking cessation mobile app and an exhaled CO checker | Low |
| **Pierce, 2020<sup>44</sup>** | USA | March - April 2020 | Analytical cross-sectional study | Primary care patients (mean age 45) at an academic family medicine centre | 6,984 | Assessed associations between sociodemographic characteristics and visit modality: | Family medicine consultations | High |
|  |  |  |  |  |  | RC (audio or video)<br>F2F |  |  |
| **Quinton, 2021<sup>45</sup>** | USA | March 2019 - March 2021 | Analytical cross-sectional study | Patients aged >18 presenting for ambulatory visits at a rural healthcare provider | 54,559 | Assessed associations between patient characteristics and visit modality:<br>RC (audio or video)<br>F2F | Ambulatory care visits | Moderate |
| **Reed, 2020<sup>46</sup>** | USA | January 2016 - May 2018 | Analytical cross-sectional study | Primary care patients of all ages at a large integrated health care delivery system | 1,131,722 | Assessed associations between patient characteristics and visit modality:<br>RC (telephone or video)<br>F2F | Index primary care consultations (no visits within 7 days prior) | Moderate |
| **Reed, 2021<sup>38</sup>** | USA | January 2016 - May 2018 | Retrospective cohort study | Primary care patients of all ages (mean age 43) at a large integrated health care delivery system | 1,131,722 | Comparison of outcomes between consultation modalities:<br>RC (telephone or video)<br>F2F | Index primary care consultations (no visits within 7 days prior) | Moderate |
| **Rene, 2022<sup>39</sup>** | USA | October-2019 - May 2020 | Retrospective cohort study | Primary care patients aged >18 with depression and/or anxiety | 338 (RC: 181; F2F: 157) | Comparison of outcomes between consultation modalities:<br>RC (pandemic | Initial behavioural health consultation, followed by 30- | Moderate |
|  |  |  |  |  |  | cohort: Apr – May 20)<br>▪ F2F (pre-pandemic cohort: Oct– Nov 19) | minute visits for behavioural activation, motivational interviewing, and psycho-education |  |
| **Ryskina, 2021<sup>40</sup>** | USA | March 2020 - May 2020 | Retrospective cohort study | Primary care patients aged >65 (mean age 75.1) | 17,103 (RC: 10,311; F2F: 6,792) | Comparison of outcomes between consultation modalities:<br>▪ RC (telephone or video)<br>▪ F2F | Primary care consultations | Moderate |
| **Tan, 2020<sup>26</sup>** | Singapore | April 2019 - May 2019 | Quasi-experimental study (prospective, self-controlled) | Active military servicemen (93% aged 18-24) accessing primary care at a military medical centre | 28 | Comparison of outcomes between consultation modalities:<br>▪ RC (on-premises Zoom video)<br>▪ F2F (immediately following RC) | Primary care consultations with assistant present to perform auscultations and physical examinations<br>Symptom collection app used prior to RC and data available to the RC physician | Moderate |
| **Ure, 2022<sup>53</sup>** | New Zealand | May - July 2021 | Cohort study | Primary care patients of all ages (25% aged <5) at a general | 454 (RC: 133; F2F: 321) | Comparison of outcomes between consultation modalities: | Primary care consultations following triage by telephone | Moderate |
|  |  |  |  | practice medical centre |  | RC<br>F2F |  |  |
| **Wickstrom, 2018<sup>54</sup>** | Sweden | October 2014 - September 2016 | Cohort study | Primary care patients (RC mean age 77; F2F mean age 75) with hard-to-heal ulcers | Healing time study: 1,988 (RC: 100; F2F: 1,888)<br>Waiting time study: 200 (RC: 100; F2F: 100) | Comparison of outcomes between consultation modalities:<br>RC (Skype video)<br>F2F | Consultations involved ulcer diagnosis and discussion of treatment strategy | Moderate |

BMI, body mass index; CO, carbon monoxide; F2F, face-to-face; GP, general practitioner; NA, not applicable; RCT, randomised controlled trial; RC, remote consultation; UK, United Kingdom; USA, United States of America
<sup>a</sup> Number of index visits; number of participants not reported
<sup>b</sup> Conditions included: sinusitis, upper respiratory infection, urinary tract infection, conjunctivitis, bronchitis, pharyngitis, influenza, cough, dermatitis, digestive symptom, or ear pain
<sup>c</sup> Conditions included: sinusitis, conjunctivitis, urinary tract infection, upper respiratory infection, influenza/ pneumonia, bronchitis, dermatitis/ eczema, ear pain, digestive symptoms, and cough

**Table 2.**
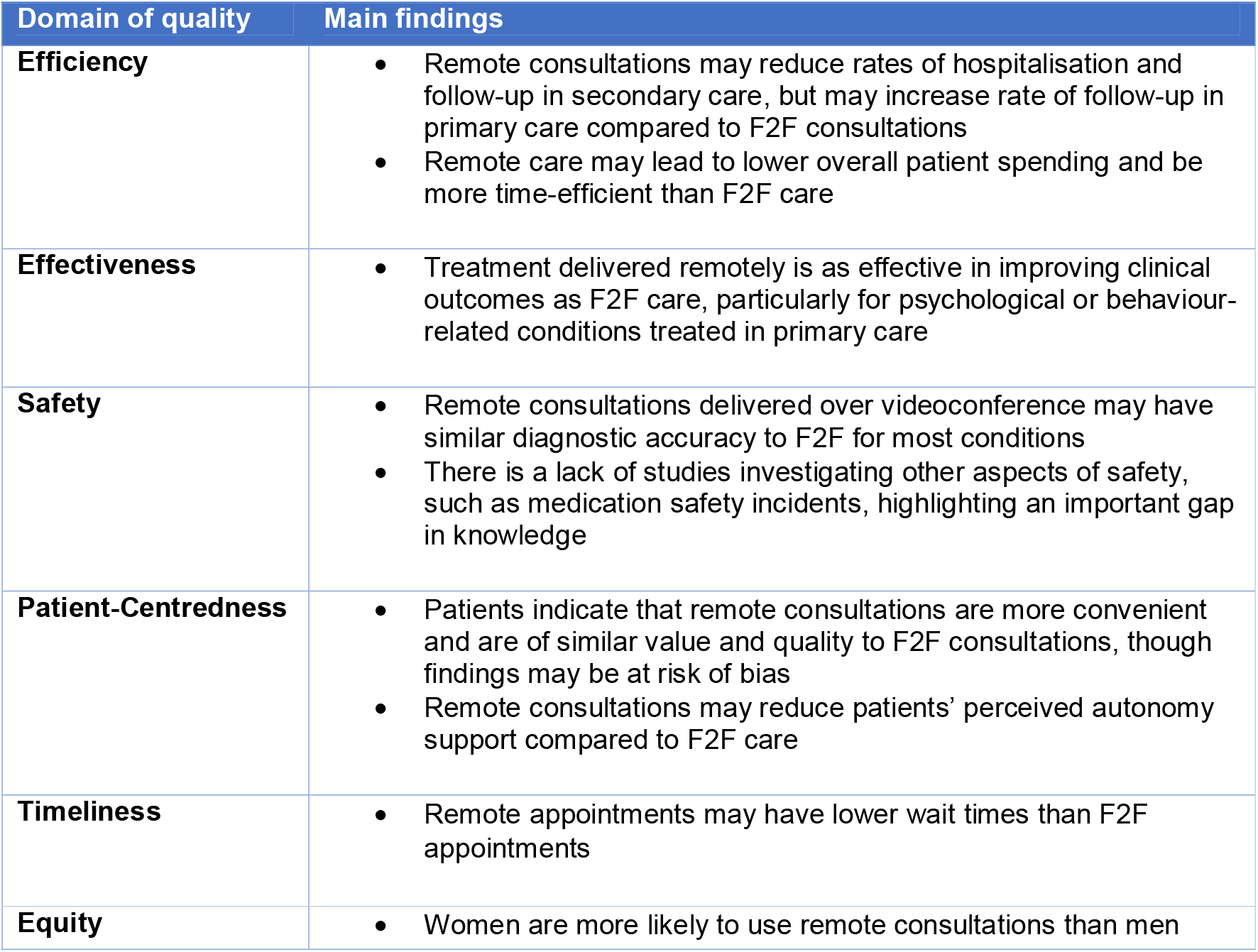
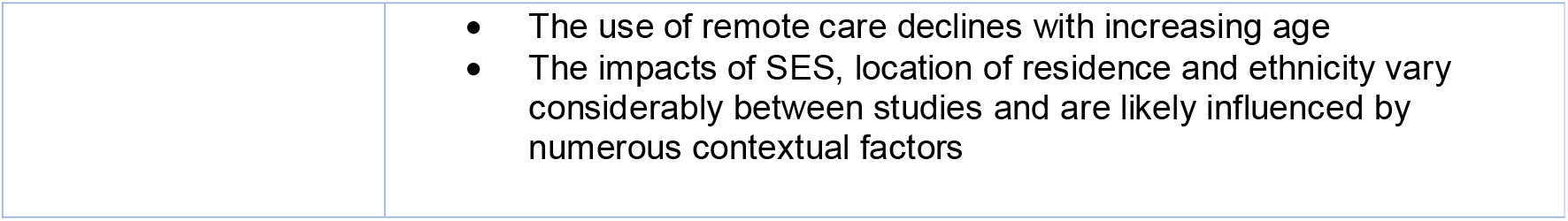
Summary of main findings.

| Domain of quality | Main findings |
| --- | --- |
| **Efficiency** | Remote consultations may reduce rates of hospitalisation and follow-up in secondary care, but may increase rate of follow-up in primary care compared to F2F consultations<br>Remote care may lead to lower overall patient spending and be more time-efficient than F2F care |
| **Effectiveness** | Treatment delivered remotely is as effective in improving clinical outcomes as F2F care, particularly for psychological or behaviour-related conditions treated in primary care |
| **Safety** | Remote consultations delivered over videoconference may have similar diagnostic accuracy to F2F for most conditions<br>There is a lack of studies investigating other aspects of safety, such as medication safety incidents, highlighting an important gap in knowledge |
| **Patient-Centredness** | Patients indicate that remote consultations are more convenient and are of similar value and quality to F2F consultations, though findings may be at risk of bias<br>Remote consultations may reduce patients' perceived autonomy support compared to F2F care |
| **Timeliness** | Remote appointments may have lower wait times than F2F appointments |
| **Equity** | Women are more likely to use remote consultations than men |
|  | • The use of remote care declines with increasing age<br>• The impacts of SES, location of residence and ethnicity vary considerably between studies and are likely influenced by numerous contextual factors |

Studies considered patients with a range of health conditions (i.e. mental illnesses,^33,37,39,50^ urgent and non-emergent conditions,^30,41^ overweight and obesity,^28,55^ low back pain,^29^ alcohol use disorders,^51^ nicotine dependence,^52^ hard-to-heal ulcers,^54^ and acute, non-urgent conditions.^34^ The remaining studies (n=17) considered primary care users in general. All consultations were delivered in primary care settings. Three studies specified occurring in rural locations^45,51,55^ and one was conducted in an urban, socioeconomically deprived area.^48^

### Summary of Risk of Bias Assessment

Nineteen studies had a moderate risk,^26,28-30,34-36,38-40,45-50,53-55^ three had a low risk,^32,37,52^ and eight had a high risk of bias^27,31,33,41-44,51^ (Figure 2). For quantitative descriptive studies, the main sources of bias were poor sample representativeness and risk of non-response bias (i.e. potential lower engagement from those with lower digital literacy).^42,43,47^ Main sources of bias for RCTs included issues with blinding,^51,52,55^ low or unclear adherence to the intervention,^50,51,55^ lack of details on randomisation,^51^ and differences between groups at baseline.^50^ Non-randomised studies had globally a high risk of bias, stemming from uncertain accuracy of measurements of exposure and outcome,^27-32,34,35,38,41,44-46^ confounders unaccounted for,^27,29,30,40,41,44,45^ overrepresentation of certain subgroups,^49^ and selection bias. ^36,38,46^

**Figure 2.**
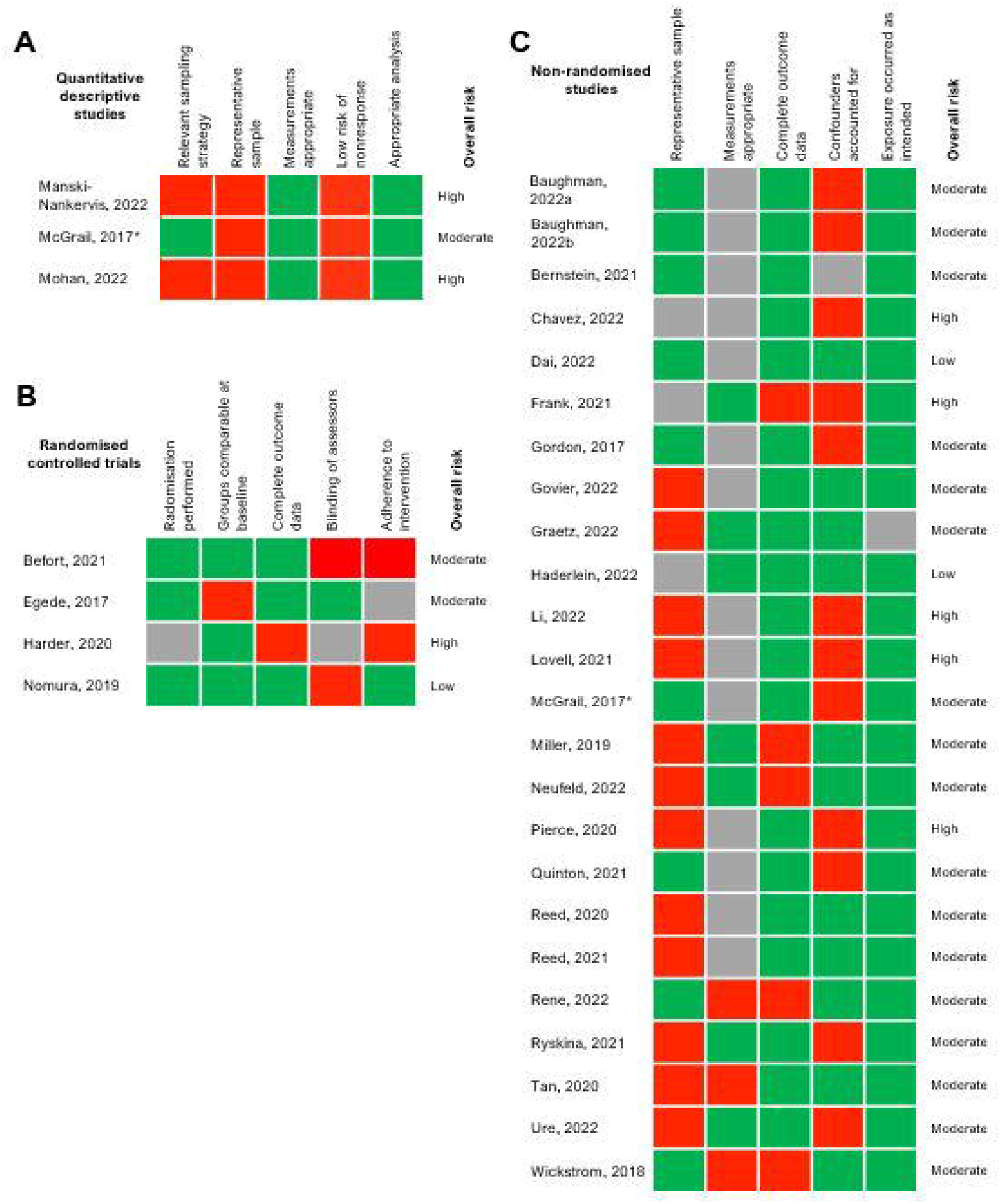
Risk of bias assessments. Cells were coloured coded green for ‘yes’, grey for ‘can’t tell’, and red for ‘no’. *McGrail is categorised as both quantitative descriptive and non-randomised due to the two distinct cross-sectional survey and quasi-experimental components of the study.

### Interventions

Four studies investigated telephone consultations only^48,49,51,55^ and ten assessed consultations delivered only over videoconference, using a range of platforms (i.e., Skype, Zoom, videophone or bespoke telehealth portals).^26,28-30,41-43,50,52,54^ The remaining sixteen studies considered both telephone and videoconference as the intervention. Almost all studies involved first consultations in an episode of care; other interventions consisted of consultations including behavioural therapy,^39,50,55^ motivational interviewing,^51^ and smoking cessation counselling.^52^

### Outcomes

#### Efficiency

Sixteen papers evaluated outcomes related to efficiency,^26,27,29-31,33,34,38-42,47,48,50,53^ including rates of follow-up visits and hospitalisations, patient costs, and appointment characteristics (i.e., length, attendance, cancellations, and no-shows) (Appendix 5). Eleven out of sixteen papers found a positive (n=6) or no impact (n=5) on efficiency in at least half of the outcomes extracted.^26,29,33,34,38,40-42,47,48,50^ Of the eight studies comparing rates of follow-up visits in primary care, five found that remote consultations resulted in a greater need for additional care,^30,31,38,41,53^ while three found no differences.^34,47,48^

Out of seven studies evaluating the rates of follow-up consultations in secondary or tertiary care, three found no changes in rates of emergency department (ED) follow-up visits or hospitalisations.^38,41,48^ Another three studies^34,40,47^ found a significant reduction in follow-up visits after a remote consultation (including ED visits,^34^ hospitalisations^34,40^ and ambulatory care sensitive conditions (ACSC) visits^40^). Only one study found that high use of remote consultations was associated with more annual ACSC visits compared to practices with low use.^27^

In terms of appointment characteristics, one study found that remote consultations led to a higher number of appointments attended and fewer cancellations.^33^ However, another reported lower rates of attendance and increased cancellations in the context of mental health appointments that took place remotely during the pandemic.^39^ One paper found that the use of remote consultation and a symptoms checker application resulted in a shorter appointment duration when compared with F2F care.^26^ Lastly, four out of the five papers assessing the impact on patient costs demonstrated a reduction in the costs associated with remote consultations compared to F2F visits.^34,41,42,47^

#### Effectiveness

Seven studies assessed the effectiveness of remote consultations (Appendix 6).^28,33,39,51,52,54,55^ Six found a non-inferior impact on effectiveness for at least half of the outcomes.^33,39,51,52,54,55^ Two studies investigating the effectiveness of remote mental health care in improving anxiety and depression symptoms also demonstrated its non-inferiority^39^ or superiority^33^ to F2F care. Remote consultations were found to be more effective for the care of hard-to-heal ulcers,^54^ and equally as effective as F2F care in reducing alcohol consumption,^51^ abstinence from smoking,^52^ and for weight management.^55^

#### Safety

Only a small study (n=28) considered outcomes related to the safety of care, finding an overall diagnostic agreement rate of 92% between remote and F2F assessments.^26^ Agreement rate was 100% for headache, gastroenteritis, and conjunctivitis, but lower for dermatological conditions and upper respiratory tract infections (87.5% and 93.3%, respectively).^26^

#### Patient-Centredness

Four of five studies assessing the impact on patient-centredness indicated that remote care had a positive or equivalent effect compared to F2F care (Appendix 7).^26,42,43,47^ One study found that those seen remotely reported lower perceived autonomy support. ^49^ Three studies asked patients to compare their recent teleconsultations with past experiences of F2F care,^42,43,47^ with most respondents agreeing that remote care was ‘as good’ (84%)^42^ and ‘as thorough’ (79%)^47^ as F2F care; more convenient (91%) and of equal or better value (67%).^43^ In another study, the majority of patients (39.9%) had no preference regarding consultation modality.^26^

#### Timeliness

Two out of the three studies evaluating the impact on timeliness found an improvement when consultations were delivered remotely (Appendix 8).^36,54^ Graetz et al^36^ reported that both video and telephone consultations were more likely to occur within one day of scheduling. Similarly, a study at a wound healing clinic found that remote consultations took place significantly sooner after referral.^54^ In contrast, a study at a Primary Care Mental Health clinic reported that patients who were initially assessed remotely were less likely to receive same-day mental health care.^37^

#### Equity

Six studies assessed the impact on equity of care in terms of the utilisation of services by sex, age, ethnicity, socio-economic status (SES) and rural or urban residence (Appendix 9).^32,35,40,44-46^ Two studies found that women were more likely to use remote care than men.^44,46^ Regarding age, two papers reported that the likelihood of having remote consultations decreased with increasing age.^32,46^ Interestingly, one study performed in the first month of the pandemic found that those over 65 were more likely to have remote consultations compared to those aged 18 to 44.^44^

Lower use of remote consultations was reported to be associated with both lower^45,46^ and higher SES.^32,44^ Notably, two studies found that those of lower SES were less likely to have video consultations than they were to have telephone consultations.^44,46^

Findings regarding the impact of ethnicity on the utilisation of remote care were also mixed. Two studies suggested that black patients were less likely to use remote consultations than white patients,^44,45^ while another three found the opposite effect.^35,40,46^ Asian patients residing in the US were more likely to use video consultations than white patients but slightly less likely to use telephone consultations.^46^

Lastly, a US study reported that patients living in rural areas had lower remote care use,^44^ whereas an Australian study found the inverse effect.^32^

## Discussion

### Summary of main findings

Our findings suggest that remote consultations are equally or more effective than F2F care for the management of conditions including mental illness, excessive smoking, and alcohol consumption. The evidence for the impact on clinical safety is extremely limited. Four studies indicated positive impacts on some aspects of patient-centredness, however, a negative impact was noted on patients’ perceived autonomy support (i.e., the degree to which people perceive others in positions of authority to be autonomy supportive). Remote consultations may reduce waiting times, lower patient costs, and reduce rates of follow-up in secondary and tertiary care. However, there is evidence that remote consultations may increase the need for additional GP visits compared with those seen in person.

Evidence regarding equity was considerably mixed. Overall, it appears that remote care is more likely to be used by younger, female patients, with disparities between other subgroups depending on contextual factors.

### Interpretation of Findings in the Context of Previous Research

#### Efficiency

The indication that remote consultations may increase rates of follow-up visits in primary care is consistent with previous evidence.^3,14^ This negative impact on efficiency may to some extent be explained by the timeframes during which the studies took place. In those occurring shortly after the onset of the pandemic,^31,53^ when the rapid transition to remote care was necessary, the increased rates of follow-up may be a consequence of lower clinician or patient confidence in remote care due to its initial unfamiliarity.^34^

Previous reviews have not specifically assessed the impact of remote consultations on follow-up at secondary or tertiary levels of care. This review’s finding that they may reduce or have no impact on follow-up at these higher levels of care might be explained by retrospective study designs precluding adequate adjustment for confounders and by the heterogeneity of the interventions included.^30,34,38,40^

The finding that remote care may be associated with lower patient costs is in line with previous research.^3,15^ Although this evidence is most relevant to countries in which patients pay for services out-of-pocket, it appears that patients accessing publicly funded health systems may also benefit financially from remote care, mainly due to reductions in travel expenses in time costs from loss of work.^3,41,42^ Furthermore, this review and wider evidence indicate that remote consultations are generally shorter than F2F visits.^3,14,26^ The decrease in consultation length reported by Tan et al^26^ may be explained by the use of a symptom checker application prior to the teleconsultation, which could have improved efficiency for clinicians. However, it is so far unclear whether shorter appointments are indeed more cost-effective than longer appointments, and whether they allow enough time for discussion of more complex matters.

#### Effectiveness

Remote care seems to be as effective as F2F visits for certain clinical outcomes (i.e., depression and anxiety symptoms, alcohol use disorder scores, smoking abstinence rates, and ulcer healing times). Existing reviews have similarly found non-inferior outcomes when remote care was delivered by specialists,^3,10^ combined with remote patient monitoring,^11^ or offered in addition to F2F care.^13^ This study suggests that remote consultations may be an effective substitute for F2F consultations in primary care settings.

#### Patient-centredness

The wider literature has reported similarly positive findings in terms of patient satisfaction across various types of secondary care and remote patient monitoring for diabetes.^16,17^ One possible explanation is that the use of remote consultations can reduce time pressure for physicians, allowing them to provide more patient-centric consultations. However, the notable positive impact on patient-centredness (i.e., convenience and preference) should be interpreted with caution, due to the significant risk of bias in these studies and the heterogeneity of measures used. Many of the included studies relied on non-validated surveys to assess satisfaction. In this review, the only study using validated questionnaires found that remote care led to lower perceived autonomy support, possibly due to the absence of non-verbal cues and decreased relational competence.^49^

#### Timeliness

There is some evidence that opting for remote care may reduce wait times for initial consultations compared to F2F care,^36,54^ possibly facilitated by removing the barriers of job flexibility and travel times.^36^ Shorter wait times is a key benefit of remote care perceived by patients.^19^ However, the finding that wait times for mental healthcare were increased following a remote appointment may reflect the possibility that patients with lower clinical need chose remote consultations to begin with, (and thus their concerns were assessed as less urgent) or indicate logistical issues in transitions of care.^37^

#### Equity

Our findings regarding age and sex are in line with previous research,^20^ and reflect older patients’ lower average digital literacy and access to technology.^44^ Evidence concerning the impacts of SES, ethnicity and location of residence on the use of remote care was inconsistent across studies, potentially due to differences in populations and study settings. To this end, access and utilisation of care is highly context-specific and will be shaped by both community and practice-level features. As engagement and participation in care is generally lowest for socially disadvantaged populations, disentangling the patient characteristics that may exclude them from remote care services, at the local level, is essential.

#### Safety

Limited conclusions can be drawn regarding the safety of remote consultations as only one study investigating outcomes in this domain was identified in this review.

### Strengths and Limitations

This study provides insight into the recent changes in the delivery of primary care and the impact of remote consultations on care quality. The review uses the IOM’s comprehensive quality framework and maps findings to this model, lending a structured approach to the evaluation of the impact of remote consultations.

This review did not consider paediatric populations, care delivered by non-healthcare professionals (*e*.*g*., community health workers), or outcomes related to medication prescribing. Many of the included studies consider patients who are part of specific subpopulations, such as elderly veterans^50^ or young military servicemen,^26^ which may limit the generalisability of the findings.

Eligible studies were restricted to those published within the last five years; while this timeframe was considered reasonable in line with the changing healthcare landscape, this might have potentially led to some earlier papers being missed.

All but one of the included studies were from high-income countries and most were from the US, highlighting the lack of research from low and middle-income countries. Many of the findings of this review will therefore have limited relevance outside of the US, and certainly outside of high-income countries with dissimilar health system financing structures or technological infrastructure. The lack of papers evaluating safety aspects, such as medication safety incidents, demonstrates the gap in knowledge previously highlighted by Gleeson et al.^18^ Similarly, current evidence has common limitations of bias introduced through lack of adjustment for confounders, and selection bias towards inclusion of patients with greater digital literacy or engagement with healthcare.

Finally, the apparent lack of studies investigating the effectiveness of remote care for a wider range of morbidities in primary care highlights a key area for further research.

### Implications for research, policy, and practice

While evidence of improved efficiency is likely to keep driving the implementation of remote care, it is critical to ensure that the transition to new service delivery models does not pose additional patient harm. The apparent lack of studies investigating the safety of remote consultations highlights a concerning gap in the literature, and future evaluations should focus on the evaluation of diagnostic error and medication safety in this context. Furthermore, health technology assessments investigating the impact on patient-centredness should capitalise on the use of validated patient-reported measures whenever possible, to allow a rigorous comparative approach.

Policy efforts to support improvements in data collection in primary care (i.e., consultation type, duration, and quality outcomes) will be critical to developing a strong evidence-base capitalising on real-world data. The mixed findings on the impact on equity highlight the need for investigations at the local level, which will be vital to develop context-specific strategies, tailored to community health needs and characteristics. Data collection should adopt an intersectional approach, considering a breadth of patient characteristics, to inform the design of locally appropriate interventions and ensure equitable access to care. Importantly, remote consultation interventions and access schemes must incorporate participatory approaches in their research and design, encouraging input from marginalised voices and including community knowledge, values, and preferences in decision-making processes.

## Supporting information

PRISMA checklist

## Data Availability

The search strategy and extracted data contributing to the narrative synthesis are available as Supplementary Data. Any additional data are available on request.

## Acknowledgments

This work is independent research supported by the National Institute for Health and Care (NIHR) Research Applied Research Collaboration Northwest London. EL and ALN are funded by NIHR Patient Safety Research Collaborative, with infrastructure support from Imperial NIHR Biomedical Research Centre. The views expressed in this publication are those of the author(s) and not necessarily those of the National Institute for Health Research or the Department of Health and Social Care.

## Contributor statement

ALN and KC contributed to the conception and design of the study. KC, EL, NO, and ALN contributed to the literature search and data extraction. KC, GG, TB, BH, and ALN contributed to data analysis and interpretation. All authors contributed to writing the manuscript and critical revision. All authors approved the manuscript. KC and ALN guarantee the integrity of the work.

## Declaration of interests

KC, GG, EL, NO, TB, AM and ALN have nothing to disclose. BH is an employee of eConsult Health Ltd, a provider of electronic consultations for NHS primary, secondary and urgent / emergency care.

