## Appendix for "The impact of remote consultations on the quality of primary care: A systematic review"

Appendices

**Appendix 1**. Medline search strategy and grey literature search.

| *Search items* | *Concepts* |
| --- | --- |
| 1 (telemedicine or tele medicine or telehealth or tele health or telecare or tele care or teleconsult* or ((virtual* or remote* or telephon* or phone* or video* or online) adj3 (consult* or appointment*))) 2 telemedicine/ or remote consultation/ 3 1 or 2 | Remote consultation |
| 4 (primary care or primary health care or primary healthcare or general practic* or general medical practice or family medicine or family practic* or family physician*) 5 exp General Practice/ 6 Primary Health Care/ 7 4 or 5 or 6 | Primary care |
| 8 (((experience* or satisfaction) adj4 (patient* or consumer* or client* or survey* or questionnaire*)) or PREM* or patient-reported experience measure* or patient-cent?red* or person-cent?red*) 9 Patient Satisfaction/ 10 8 or 9 | Patient-centredness |
| 11 Treatment Outcome/ or ((health or clinical* or treatment*) adj3 (outcome* or effective* or efficacy)) | Effectiveness |
| 12 ((patient adj3 (safety or harm)) or misdiagnos* or safety manag* or (accident* adj2 prevent*) or error* or medication reconcil* or near miss*) 13 patient harm/ or patient safety/ or Diagnostic Errors/ 14 12 or 13 | Safety |
| 15 (efficiency or economic* or cost* or expenditure* or charge* or (number adj3 appointment*) or (number adj3 admission*) or (number adj3 consultation*)) | Efficiency |
| 16 (wait* list* or wait* time* or timeliness) 17 Time-to-Treatment/ or Waiting Lists/ 18 16 or 17 | Timeliness |
| 19 ((health* or health care or access) adj3 (equity or disparit* or inequit* or inequalit* or equality or gap)) 20 Health Equity/ 21 19 or 20 | Equity |
| 22 10 or 11 or 14 or 15 or 18 or 21 23 3 and 7 and 22 |  |

**Appendix 2**. Study inclusion and exclusion criteria

|  | Inclusion criteria | Exclusion criteria |
| --- | --- | --- |
| Population (and setting) | Adult patients (mean age ≥18 years) with any health condition, accessing primary care services in any geographical location | Patients accessing secondary, tertiary, or quaternary care; direct-to-consumer services; care delivered in retail clinics; or care that is not integrated into primary care |
| Intervention | Two-way, synchronous patient-provider remote consultations delivered via:   - **telephone** - **videoconference**   by primary care healthcare professionals, or multi-component interventions involving synchronous remote consultations | Consultations involving only asynchronous communication; synchronous online messaging; remote-patient monitoring, automated services; or interventions for education or administrative purposes  Consultations delivered by non-healthcare professionals or specialist clinicians, delivered in retail clinics or by direct-to-consumer models |
| Comparator | Consultation delivered face-to-face, or the outcomes assessed indicate comparison with previous experience of face-to-face care (survey questions) | No face-to-face comparison group; or no indication of comparison with face-to-face care |
| Outcomes | Studies reporting any quantitative measures related to (1) efficiency (e.g., service costs, follow-up care), (2) effectiveness (e.g., health outcomes); (3) patient safety (e.g., misdiagnoses); (4) patient-centredness (e.g., patient satisfaction measures), (5) timeliness (e.g., wait times) and (6) equity (e.g., disparities in access or outcomes between different patient subgroups) | Studies reporting only qualitative outcomes; outcomes that do not fit under any of the IOM’s quality framework domains  Studies evaluating prescribing outcomes only, as changes in prescription patterns are not necessarily reflective of the quality of care and are highly context specific |
| Study type | Randomised controlled trials, cluster randomised trials, quasi-experimental studies, case-control studies, cohort studies, cross-sectional studies, cost-effectiveness studies | Incomplete studies, interim reports, scoping reviews, case series, case reports, opinion pieces, trial protocols |

**Appendix 3**. Data extraction form template

| Author, year |
| --- |
| Country of publication |
| Study type |
| Sample size (n (i; c)) |
| Date of intervention |
| Duration of intervention |
| Participants |
| Setting |
| Source of data |
| Study design and comparison |
| Retention or adherence rate (n (i; c)) |
| Response rate (n (i; c)) |
| Consultation description |
| Type of technology |
| Outcomes assessed |
| Key results (mean difference, OR, RRR) |
| Adjustment for confounders |
| Domain(s) of quality |
| Method of recruitment |
| Source of funding |
| Possible conflicts of interest |

**Appendix 4**. Mixed Methods Appraisal Tool assessments with justifications for decisions.

| QUANTITATIVE DESCRIPTIVE STUDIES | | | | | | |
| --- | --- | --- | --- | --- | --- | --- |
| Author, year | Is the sampling strategy relevant to address the research question? | Is the sample representative of the target population? | Are the measurements appropriate? | Is the risk of nonresponse bias low? | Is the statistical analysis appropriate to answer the research question? | Comments |
| Manski-Nankervis, 2022^42^ | No | No | Yes | No | Yes | Sampling strategy (online surveys) may have led to a selection bias Sample was not representative of target population (highly educated, young and majority female)  Low response rate likely to introduce bias |
| McGrail, 2017^47^ | Yes | No | Yes | No | Yes | Sampling strategy (online surveys) may have led to a selection bias  Low response rate likely to introduce bias  Survey sample was not representative of entire target population (majority female and married)  No matching for race and ethnicity which may confound results |
| Mohan, 2022^43^ | No | No | Yes | No | Yes | 26.7% response rate may have introduced bias and selected for those with better digital literacy Overrepresentation of participants who were female and from higher educational/income backgrounds  Study only captures those who could attend a remote visit |
| RANDOMISED CONTROLLED TRIALS | | | | | | |
| Author, year | Is randomization appropriately performed? | Are the groups comparable at baseline? | Are there complete outcome data? | Are outcome assessors blinded to the intervention provided? | Did the participants adhere to the assigned intervention? | Comments |
| Befort, 2021^55^ | Yes | Yes | Yes | No | No | No blinding of outcome assessors to the intervention  Session attendance was notably different between the groups from 6 - 24 months |
| Egede, 2017^50^ | Yes | No | Yes | Yes | Can't tell | There were significant differences between groups in health status compared to the previous year (*P* = 0.02) Unclear if there were differences between treatment groups in terms of completion of interventions |
| Harder, 2020^51^ | Can't tell | Yes | No | Can't tell | No | No description of how randomisation was performed23% of those initially randomised did not complete intervention  No mention of assessors being blinded to the interventions. |
| Nomura, 2019^52^ | Yes | Yes | Yes | No | Yes | Outcome assessors were not blinded to intervention assignment |
| NON-RANDOMISED STUDIES | | | | | | |
| Author, year | Are the participants representative of the target population? | Are measurements appropriate regarding both the outcome and intervention (or exposure)? | Are there complete outcome data? | Are the confounders accounted for in the design and analysis? | During the study period, is the intervention administered (or exposure occurred) as intended? | Comments |
| Baughman, 2022a^28^ | Yes | Can't tell | Yes | No | Yes | Possible in accuracies in recording visit modality  Considerable variability of eligible follow-up plans which may have taken varying lengths of time to complete  Confounders such as number of visits or type of follow up plan were not accounted for |
| Baughman, 2022b^29^ | Yes | Can't tell | Can't tell | No | Can't tell | Possible in accuracies in recording visit modality and identifying patients with red flag complaints  Confounders such as severity of pain not accounted for |
| Bernstein, 2021^30^ | Yes | Can't tell | Yes | Can't tell | Yes | Possible in accuracies in recording visit modality  Possible misclassification of resolved episodes due to 30-day cut off window  Confounders mostly accounted for except for severity of condition |
| Chavez, 2022^31^ | Can't tell | Can't tell | Yes | No | Yes | No data on patient characteristics other than mean age and sex  Possible in accuracies in recording visit modality  Possible misclassification of short interval-follow ups due to 60-day cut off window |
| Dai, 2022^32^ | Yes | Can't tell | Yes | Yes | Yes | Possible in accuracies in recording visit modality  Possible misclassification of pension status |
| Frank, 2021^33^ | Can't tell | Yes | No | No | Yes | Missing data for outcomes on number of psychiatric problems  Confounders not accounted for  Small sample (n = 18) for outcomes on clinical effectiveness  Academic centre may limit generalisability to wider population |
| Gordon, 2017^34^ | Yes | Can't tell | Yes | No | Yes | Possible in accuracies in recording visit modality |
| Govier, 2022^35^ | No | No | Yes | Yes | Yes | Includes only patients who tested positive for COVID-19, may have missed those who did not test and may be biased towards those with better access to testing May not have captured care accessed by patients in a different healthcare system |
| Graetz, 2022^36^ | No | Yes | Yes | Yes | Can't tell | Sample may not be representative of target population as only included appointments booked via online portal  Unclear if the appointments actually occurred by the modality requested by the patient |
| Haderlein, 2022^37^ | Can't tell | Yes | Yes | Yes | Yes | Sample may not be representative of the wider target population |
| Li, 2022^27^ | No | Can't tell | Yes | No | Yes | May not be wholly representative of the target population Potential inaccuracies of claims data  Possible confounders are not accounted for as data is unknown |
| Lovell, 2021^41^ | No | Can't tell | Yes | No | Yes | May not be wholly representative of the target population  Possible confounders not accounted for as data is unknown |
| McGrail, 2017^47^ | Yes | Can't tell | Yes | No | Yes | Sampling strategy (online surveys) may have led to a selection bias Low response rate likely to introduce bias  Survey sample was not representative of entire target population (majority female and married)  No matching for race and ethnicity which may confound results |
| Miller, 2019^48^ | No | Yes | No | Yes | Yes | May not be generalisable to wider population  Did not report any numerical evidence for change in delays to appointment |
| Neufeld, 2022^49^ | No | Yes | No | Yes | Yes | Sample not representative of wider population (middle-to-upper class, majority white and female)  Use of convenience sampling at the physician's discretion may also have led to some selection bias  Survey response rate was 63.5% |
| Pierce, 2020^44^ | No | Can't tell | Yes | No | Yes | Sample may not be representative of wider population  Potential inaccuracies of claims data Possible confounders not accounted for |
| Quinton, 2021^45^ | Yes | Can't tell | Yes | No | Yes | Potential inaccuracies of data  Possible confounders not accounted for |
| Reed, 2020^46^ | No | Can't tell | Yes | Yes | Yes | Sample may not be representative of wider population Potential inaccuracies of data and misclassification of patients in sociodemographic groups (e.g. socioeconomic status was inferred from area level data). |
| Reed, 2021^38^ | No | Can't tell | Yes | Yes | Yes | Sample may not be representative of wider population Potential inaccuracies of data and misclassification of index visits or patients requiring follow-ups due to the 7 day time frame used |
| Rene, 2022^39^ | Yes | No | No | Yes | Yes | Potential inaccuracies of data  Unclear follow-up period |
| Ryskina, 2021^40^ | No | Yes | Yes | No | Yes | Hispanic patients were underrepresented in the sample  Possible confounders not accounted for |
| Tan, 2020^26^ | No | No | Yes | Yes | Yes | Sample very small and not representative of wider military population  Did not use a validated satisfaction questionnaire |
| Ure, 2022^53^ | No | Yes | Yes | No | Yes | Not representative of wider population  Possible confounders not accounted for |
| Wickstrom, 2018^54^ | Yes | No | No | Yes | Yes | Use of different measurement techniques between the study and control groups may have resulted in differences between groups  Low rates of 6 month follow up, especially for the control group |

**Appendix 5*.*** The efficiency of remote vs face-to face (F2F) consultations.

| *Author, year* | *Outcome measure* | *RC mean (95% CI or SD)* | *F2F mean (95% CI or SD)* | *Mean difference or effect size (95% CI and/or P value)* | *Risk of bias* |
| --- | --- | --- | --- | --- | --- |
|  | **a) Rates of follow up and hospitalizations** | | |  |  |
| Bernstein, 2021^30^ | Visit resolution (no follow-up within 30 days) (%) | 84.00 (76.50, 90.90) | 90.70 (87.70, 93.40) |  | Moderate |
|  | Episodes of care required for resolution | 2.50 (2.11, 2.72) | 2.90 (2.01, 2.42) |  |  |
|  | Index visits requiring additional episodes of care (%) | 14.80 | 8.10 |  |  |
| Chavez, 2022^31^ | Short-interval follow-up rate (any visit within 60 days) (%) | 22.80 | 16.80 | (*P* < 0.001) | High |
|  | Visits requiring follow-up within 15 days (%) | 31.34 | 23.44 | (*P* < 0.001) |  |
| Li, 2022^27^ | Rate of ACSC visits^a^ (high practice RC vs low practice RC) |  |  | 2.10 (0.22, 3.97) | High |
|  | Rate of ACSC visits^a^ (medium practice RC vs low practice RC) |  |  | 0.69 (-0.93, 2.21) |  |
| Lovell, 2021^41^ | ED follow-up rate (within 21 days) (%) | 1.80 | 2.60 | 1.49 (0.87, 2.12;*P* > 0.05) | High |
|  | Inpatient follow-up rate (within 21 days) (%) | 0.40 | 0.70 | 1.77 (0.22, 3,32;P > 0.05) |  |
|  | Any visit follow-up rate (within 21 days) (%) | 35.30 | 35.70 | 1.01 (0.94, 1.09:*P* > 0.05) |  |
|  | E&M follow-up rate (within 21 days) (%) | 26.60 | 22.60 | 0.85 (0.77, 0.93;*P* < 0.001) |  |
| McGrail, 2017^47^ | GP follow-up rate (within 30 days) (%) | 1.55 | 1.43 | (*P* = 0.45) | Moderate |
|  | Medical specialist follow-up rate (within 30 days) (%) | 1.72 | 1.62 | (*P* = 0.58) |  |
|  | Surgical specialist follow-up rate (within 30 days) (%) | 1.07 | 2.18 | (*P* < 0.001) |  |
| Gordon, 2017^34^ | Outpatient follow-up rate (within 3 weeks) (%) | 28.09 | 28.10 | (*P* = 0.96) | Moderate |
|  | ED follow-up rate (within 3 weeks) (%) | 1.32 | 1.84 | (*P* = 0.02) |  |
|  | Inpatient hospitalization (within 3 weeks) (%) | 0.15 | 0.37 | (*P* = 0.02) |  |
| Reed, 2021^38^ | Office follow-up rate (within 7 days) (%) (RC = video) | 25.40 (24.70, 26.00) | 24.50 (24.50, 24.60) | (*P* < 0.05) | Moderate |
|  | Office follow-up rate (within 7 days) (%) (RC = telephone) | 26.00 (25.90, 26.20) | 24.50 (24.50, 24.60) | (*P* < 0.05) |  |
|  | ED follow-up rate (within 7 days) (%) (RC = video) | 1.23 (1.06, 1.40) | 1.30 (1.29, 1.32) | (*P* > 0.05) |  |
|  | ED follow-up rate (within 7 days) (%) (RC: telephone) | 1.37 (1.33, 1.41) | 1.30 (1.29, 1.32) | (*P* > 0.05) |  |
|  | Hospitalization rate (within 7 days) (%) (RC = video) | 0.23 (0.14, 0.32) | 0.23 (0.22, 0.24) | (*P* > 0.05) |  |
|  | Hospitalization rate (within 7 days) (%) (RC = telephone) | 0.22 (0.21, 0.24) | 0.23 (0.22, 0.24) | (*P* > 0.05) |  |
| Ure, 2022^53^ | Rate of re-triage (within 7 days) (%) | 14.00 | 7.00 | (P < 0.05) | Moderate |
| Ryskina, 2021^40^ | Odds of ACSC hospitalization (within 14 days) (RC vs F2F) |  |  | 0.78 (0.61, 1.00; *P* = 0.049) | Moderate |
|  | Odds of all-cause hospitalization (within 14 days) (RC vs F2F) |  |  | 0.72 (0.57, 0.90; *P* = 0.004) |  |
| Miller, 2019^48^ | Number of GP visits per patient per year |  |  | (*P* = 0.193) | Moderate |
|  | Number of referrals to hospital per month | 171 | 182 | 11 (*P* = 0.181) |  |
|  | Number of visits to GP out-of-hours per month | 313 | 304 | 9(*P* = 0.56) |  |
|  | Sum of local ED visits per intervention period | 3700 of 66228 | 6771 of 122428 | (*P* = 0.73) |  |
|  | **b) Patient costs** |  |  |  |  |
| Gordon, 2017^34^ | Cost of index visit (USD) | $49 | $109 | $60 (*P* < 0.001) | Moderate |
|  | Average medical costs within 3-week follow-up (USD) | $200 | $288 | (*P* < 0.001) |  |
| Lovell, 2021^41^ | Cost of index visit (USD) | $45 | $114 | 2.54 (2.46, 2.62;*P* < 0.001) | High |
|  | Pharmacy costs within 21 days of index (USD) | $111 | $117 | 1.06 (0.82, 1.29; *P* > 0.05) |  |
|  | Follow-up costs within 21 days (USD) | $288 | $490 | 1.70 (1.04, 2.36;*P* = 0.038) |  |
|  | Average total costs (USD) | $429 | $707 | 1.65 (1.26, 2.04;*P* < 0.001) |  |
| Egede, 2017^50^ | Inpatient cost trajectories over time^b^ (RC vs F2F) |  |  | 0.17 (*P* = 0.845) | Moderate |
|  | Outpatient cost trajectories over time^b^ (RC vs F2F) |  |  | −0.07 (*P* = 0.071) |  |
|  | Pharmacy cost trajectories over time^b^ (RC vs F2F) |  |  | 0.01 (*P* = 0.805) |  |
| Manski-Nankervis, 2022^42^ | Mean total cost saving^c^ from RC vs F2F (AUD) |  |  | $61.36 ($53.24, $69.48) | High |
| McGrail, 2017^47^ | Trend in cost of primary care services^d^ (CAD) (RC vs F2F) |  |  | -$3.79 (*P* = 0.01) | Moderate |
|  | **c) Appointment characteristics** | |  |  |  |
| Frank, 2021^33^ | Number of appointments attended | 2.17 (4.46) | 1.19 (2.08) | (*P* = 0.002) | High |
|  | Number of appointment cancellations | 0.14 (0.49) | 0.53 (1.03) | (*P* < 0.001) |  |
| Rene, 2022^39^ | Number of appointment cancellations | 0.45 (0.81) | 0.36 (0.76) | (*P* = 0.003) | Moderate |
|  | Number of appointment no-shows | 0.38 (0.67) | 0.25 (0.54) | (*P* = 0.26) |  |
|  | Number of appointments attended | 2.15 (2.24) | 3.32 (1.49) | (*P* < 0.001) |  |
| Tan, 2020^26^ | Consultation length | 6 min 19 sec | 8 min 34 sec | (*P* = 0.048) | Moderate |
| Baughman, 2022b^29^ | Patients receiving imaging within 28 days (%) | 11.20 | 16.32 | 5.12(*P* < 0.01) | Moderate |

ACSC, ambulatory care-sensitive conditions; CI, confidence intervals; ED, emergency department; E&M, evaluation and management; F2F, face-to-face; GP, general practitioner; RC, remote consultation; SD, standard deviation

^a^ Visits per 1000 patients per year

^b^ Including the 8-week intervention period, and the two years prior- and post-intervention

^c^ Including paid work, unpaid time and travel costs

^d^ Average change in spending per quarter over 3 years

**Appendix 6**. The effectiveness of remote vs face-to-face (F2F) consultations.

| *Author, year* | *Outcome measure* | *RC mean (95% CI or SD)* | *F2F mean (95% CI or SD)* | *Mean difference (95% CI and/or P value)* | *Risk of bias* |
| --- | --- | --- | --- | --- | --- |
| Baughman, 2022a^28^ | Patients with completed weight management plan at 1.5 years (%) | 7.90 | 12.19 | 4.29(2.84, 5.54: *P* < 0.001) | Moderate |
|  | Patients with completed weight management plan at 3 months (%) | 1.80 | 6.59 | 4.79(3.99, 5.35;P < 0.001) |  |
|  | Patients with completed weight management plan at 1.5 years (%) (blended RC vs F2F) | 24.82 | 12.19 | 12.65%(12.29, 13.01;P < 0.001) |  |
|  | Patients with completed weight management plan at 3 months (%) (blended RC vs F2F) | 17.75 | 6.59 | 11.16 (10.85, 11.48;P < 0.001) |  |
| Befort, 2021^55^ | Weight loss (kgs) at 24 months (RC vs group F2F) | –3.90 (–5.00, –2.90) | –4.40 (–5.50, –3.40) | –0.50 ( –1.90, 0.90;*P* = 0.48) | Moderate |
|  | Weight loss (kgs) at 24 months (RC vs individual F2F) | –3.90 (–5.00, –2.90) | –2.60 (–3.60, –1.50) | –1.40 (–3.00, 0.30;*P* = .06) |  |
| Frank, 2021^33^ | CGI-S scores at end of 8-month wave | 3.33 (0.97) | 3.61(0.70) | (*P* = 0.02) | High |
|  | CGI-I scores at end of 8-month wave | 2.44 (0.51) | 3.06 (0.87) | (*P* = 0.002) |  |
| Rene, 2022^39^ | Change in PHQ-9 score | -2.90 | -2.80 | (*P* > 0.05) | Moderate |
|  | Change in GAD-7 score | -3.10 | -2.30 | (*P* > 0.05 |  |
| Harder, 2020^51^ | AUDIT-C scores at 1 month follow-up |  |  | 0.20(-0.60, 1.00;*P* = 0.63) | High |
|  | AUDIT-C scores at 6 months follow-up |  |  | 0.44(-0.47, 1.36; *P* = 0.34) |  |
| Nomura, 2019^52^ | CAR (%) from weeks 9-12 | 81.00 (71.00, 91.00) | 78.90 (68.00, 89.00) | 2.10 (–12.80, 17.00) | Low |
|  | CAR (%) from weeks 9 -24 | 74.10 (63.00, 85.00) | 71.90 (60.00, 84.00) | 2.20 (–14.00, 18.40) |  |
| Wickstrom, 2018^54^ | Number of days between consultation and complete  ulcer healing | 78 (40, 78) | 118 (75, 89) | (*P* < 0.001) | Moderate |

AUDIT-C, Alcohol Use Disorders Identification Test – Consumption; CAR, continuous abstinence rate; CGI-I, Clinical Global Impressions – Improvement; CGI-S, Clinical Global Impressions – Severity; CI, confidence intervals; F2F, face-to-face; GAD-7, Generalised Anxiety Disorder-7; kgs, kilograms; PHQ-9, Patient Health Questionnaire-9; RC, remote consultation; SD, standard deviation

**Appendix 7.** The impact of remote consultations on patient-centredness.

| *Author, year* | *Outcome measure* | *RC mean (SE)* | *F2F mean (SE)* | *Mean difference (P value)* | *Risk of bias* |
| --- | --- | --- | --- | --- | --- |
| Neufeld, 2022^49^ | Patient perceived autonomy support^a^ | 5.75 (0.17) | 6.28 (0.16) | (*P* = 0.032) | Moderate |
|  | **Survey question responses** | | |  |  |
|  |  | ***Agree/ Better*** | ***Neutral/ Equal*** | ***Disagree/ Worse*** | ***Risk of bias*** |
| Manski-Nankervis, 2022^42^ | RC visit was as good as F2F (%) | 84.00 |  | 11.40 | High |
| McGrail, 2017^47^ | RC visit was as thorough as F2F (%) | 79.00 |  | 21.00 | Moderate |
| Mohan, 2022^43^ | Convenience of RC compared to F2F (%) | 91.00 | 4.00 | 4.00 | High |
|  | Value of RC compared to F2F (%) | 37.00 | 30.00 | 33.00 |  |
| Tan, 2020^26^ | F2F visits provide better quality care than RC (%) | 32.00 | 50.10 | 17.90 | Moderate |
|  | Prefer F2F visits over RC in the future (%) | 28.60 | 39.90 | 32.10 |  |

F2F, face-to-face; RC, remote consultation; SE, standard error

^a^ Measured using the Healthcare Climate and Basic Need Satisfaction in Relationships questionnaires

**Appendix 8.** The timeliness of remote vs face-to-face consultations.

| *Author, year* | *Outcome measure* | *RC mean (95% CI or SD)* | *F2F mean (95% CI or SD)* | *Mean difference (P value)* | *Risk of bias* |
| --- | --- | --- | --- | --- | --- |
| Graetz, 2022^36^ | Days between scheduling and visit (RC = telephone) | 1.80 | 3.50 | 1.70 | Moderate |
|  | Days between scheduling and visit (RC = video) | 2.30 | 3.50 | 1.20 |  |
|  | Visits occurring within 1 day of scheduling (%) (RC: telephone) | 66.60 (66.40, 66.80) | 46.50 (46.40, 46.60) |  |  |
|  | Visits occurring within 1 day of scheduling (%) (RC: video) | 56.60 (55.90, 57.30) | 46.50 (46.40, 46.60) |  |  |
| Haderlein, 2022^37^ | Patients receiving same-day mental health care (%) | 19.70 | 36.00 | (*P* < 0.01) | Low |
| Wickstrom, 2018^54^ | Number of days between referral and consultation | 25.00 | 43.00 | (*P* = 0.017) | Moderate |

CI, confidence intervals; F2F, face-to-face; RC, remote consultation; SD, standard deviation

**Appendix 9**. The impact of remote consultations on equity of care.

| *Author, year* | *Outcome measure* | *Mean difference (%), OR, or RRR* | *95% CI and/or P value* | *Risk of bias* |
| --- | --- | --- | --- | --- |
|  | **a) Use of RC vs F2F** |  |  |  |
| Dai, 2022^32^ | Male vs female | 1.02 | 1.0, 1.04 | Low |
|  | 70-74 vs 65-69 | 0.96 | 0.9, 1.03 |  |
|  | 75-79 vs 64-69 | 0.87 | 0.82, 0.93* |  |
|  | 80-84 vs 65-69 | 0.86 | 0.81, 0.91* |  |
|  | 85+ vs 64-69 | 0.70 | 0.66, 0.74* |  |
|  | Pension holders^a^ vs not on pension | 1.14 | 1.10, 1.17* |  |
|  | Rural vs urban | 1.72 | 1.57, 1.90* |  |
| Pierce, 2020^44^ | Female vs male | 1.15 | 1.04, 1.26* | High |
|  | <18 vs 18-44 | 0.35 | 0.29, 0.41* |  |
|  | 45-65 vs 18-44 | 1.08 | 0.97, 1.21 |  |
|  | >65 vs 18-44 | 1.21 | 1.05, 1.40* |  |
|  | Black vs white | 0.65 | 0.56, 0.75* |  |
|  | Other race vs white | 0.64 | 0.50, 0.82* |  |
|  | Hispanic vs non-Hispanic | 0.87 | 0.62, 1.22 |  |
|  | Self-pay vs insurance | 1.26 | 1.04, 1.52* |  |
|  | Medicaid vs insurance | 1.29 | 1.04, 1.61* |  |
|  | Medicare vs insurance | 1.37 | 1.18, 1.60* |  |
|  | Rural vs urban | 0.81 | 0.74, 0.90* |  |
| Ryskina, 2021^40^ | Female vs male | 1.11 | 0.99, 1.59 | Moderate |
|  | 75-84 vs 65-74 | 1.1 | 0.97, 1.24 |  |
|  | >84 vs 65-74 | 1.18 | 1.0, 1.41 |  |
|  | Black vs white | 1.30 | 1.14, 1.47* |  |
|  | Asian vs white | 0.73 | 0.47, 1.12 |  |
|  | **b) Use of RC vs no care** |  |  |  |
| Govier, 2022^35^ | Non-Hispanic Black vs non-Hispanic white | + 3.30 | *P* < 0.05* | Moderate |
|  | Non-Hispanic Asian vs non-Hispanic white | + 0.30 | *P* > 0.05 |  |
|  | Non-Hispanic NH/PI vs non-Hispanic white | + 0.60 | *P* > 0.05 |  |
|  | Non-Hispanic other vs non-Hispanic white | − 2.53 | *P* < 0.05* |  |
|  | Hispanic/Latino vs non-Hispanic white | − 2.11 | *P* < 0.01* |  |
|  | SES Theme^b^ | + 0.03 | *P* > 0.05 |  |
|  | Minority Status/Language Theme^b^ | − 0.83 | *P* > 0.05 |  |
| Quinton, 2021^45^ | Female vs male | 0.93 | 0.80, 1.08 | Moderate |
|  | Black vs white | 0.54 | 0.42, 0.70; *P* < 0.05* |  |
|  | Other vs white | 0.86 | 0.44, 1.67 |  |
|  | Self-pay vs commercial | 0.73 | 0.55, 0.96; *P* < 0.05* |  |
|  | Medicaid vs commercial | 0.44 | 0.31, 0.61; *P* < 0.05* |  |
|  | Medicare vs commercial | 0.70 | 0.55, 0.89; *P* < 0.05* |  |
|  | 80-100% broadband access vs 0-20% | 2.24 | 1.70, 2.95; *P* < 0.05* |  |
|  | 60-80% broadband access vs 0-20% | 1.10 | 0.86, 1.41; *P* < 0.05* |  |
|  | 40-60% broadband access vs 0-20% | 1.07 | 0.54, 2.10; *P* < 0.05* |  |
|  | 20-40% broadband access vs 0-20% | 1.09 | 0.86, 1.37 |  |
|  | Partially rural vs rural | 2.64 | 2.25, 3.11; *P* < 0.05* |  |
|  | Non-rural vs rural | 0.69 | 0.26, 1.88 |  |
|  | **c) Use of video vs F2F** |  |  |  |
| Reed, 2020^46^ | Male vs female | 0.93 | 0.90, 0.96* | Moderate |
|  | <18 vs 18-44 | 1.00 | 0.91, 1.09 |  |
|  | 45-64 vs 18-44 | 0.61 | 0.58, 0.63* |  |
|  | >65 vs 18-44 | 0.24 | 0.22, 0.26* |  |
|  | Black vs white | 1.62 | 1.52, 1.73* |  |
|  | Hispanic vs white | 0.92 | 0.88, 0.97* |  |
|  | Asian vs white | 1.26 | 1.22, 1.32* |  |
|  | Low SES neighbourhood vs high | 0.93 | 0.89, 0.97* |  |
|  | **d) Use of telephone vs F2F** |  |  |  |
| Reed, 2020^46^ | Male vs female | 0.80 | 0.79, 0.81* | Moderate |
|  | <18 vs 18-44 | 0.42 | 0.40, 0.43* |  |
|  | 45-64 vs 18-44 | 0.80 | 0.70, 0.81* |  |
|  | >65 vs 18-44 | 0.55 | 0.54, 0.57* |  |
|  | Black vs white | 1.28 | 1.25, 1.31* |  |
|  | Hispanic vs white | 1.00 | 0.98, 1.01 |  |
|  | Asian vs white | 0.96 | 0.94, 0.97* |  |
|  | Low SES neighbourhood vs high | 1.01 | 1.00, 1.03 |  |
|  | **e) Use of video vs telephone** |  |  |  |
| Dai, 2022^32^ | Male vs female | 0.94 | 0.86, 1.02 | Low |
|  | 70-74 vs 65-69 | 0.80 | 0.61, 1.03 |  |
|  | 75-79 vs 64-69 | 1.13 | 0.89, 1.44 |  |
|  | 80-84 vs 65-69 | 1.03 | 0.82, 1.29 |  |
|  | 85+ vs 64-69 | 1.20 | 0.97, 1.49 |  |
|  | Pension holders^a^ vs not on pension | 1.01 | 0.90, 1.13 |  |
|  | Rural vs urban | 0.41 | 0.29, 0.57* |  |
| Pierce, 2020^44^ | Female vs male | 1.08 | 0.91, 1.27 | High |
|  | <18 vs 18-44 | 0.92 | 0.59, 1.45 |  |
|  | 45-65 vs 18-44 | 0.51 | 0.41, 0.62* |  |
|  | >65 vs 18-44 | 0.27 | 0.21, 0.33* |  |
|  | Black vs white | 0.72 | 0.55, 0.93* |  |
|  | Other race vs white | 0.96 | 0.48, 1.82 |  |
|  | Hispanic vs non-Hispanic | 0.93 | 0.48, 1.82 |  |
|  | Self-pay vs insurance | 0.68 | 0.49, 0.95* |  |
|  | Medicaid vs insurance | 0.36 | 0.26, 0.51* |  |
|  | Medicare vs insurance | 0.79 | 0.64, 0.99* |  |
|  | Rural vs urban | 1.36 | 1.14, 1.61* |  |

CI, confidence intervals; F2F, face-to-face; NH/PI, Native Hawaiian/ Pacific Islander; OR, odds ratio; RC, remote consultation; RRR, relative risk ratio; SES, socioeconomic status

* Statistically significant findings

^a^ Pension holder status was used as a proxy indicator of lower SES

^b^ Reference category indicates those less vulnerable for the respective theme, defined using the Social Vulnerability Index
